# Evaluating Cost-effectiveness of 9-valent HPV Vaccination for Men Who Have Sex with Men by HIV Status in Hong Kong

**DOI:** 10.1101/2025.06.02.25328835

**Authors:** Dijing You, Jianchao Quan, David Bishai, Wendy Wing Tak Lam, Karen Ann Grépin, Linda Chan, Diana Dan Wu, David Ka Ki Wong, William Chi Wai Wong, Jiandong Zhou

## Abstract

**Background:** Human papillomavirus (HPV) is the most common sexually transmitted infection and a leading cause of anal cancer and genital warts, particularly among men who have sex with men (MSM). In Hong Kong, HPV vaccination is currently only offered to school-aged girls, despite the high burden of HPV-related diseases among MSM, especially those living with HIV. Existing cost-effectiveness evaluations of HPV vaccination in Hong Kong primarily focus on female-only strategies and heterosexual men without accounting for differences in HPV infection risks among MSM. In contrast, countries such as the United Kingdom initially implemented free HPV vaccination for MSM through sexual health clinics, followed by an expansion of the programme to include adolescent boys. This provides a potentially useful model for Hong Kong to consider. This study aimed to evaluate the cost-effectiveness of implementing 9-valent HPV (9vHPV) vaccination strategies among HIV-positive and HIV-negative MSM in Hong Kong.

**Methods:** We developed a Markov model to simulate the natural history of HPV infection and progression to genital warts, and anal cancer in a cohort of 100,000 MSM in Hong Kong, stratified by four age groups (12–18, 19–27, 28–45, and >45 years) and HIV status (positive or negative). A 10-year time horizon was used from the healthcare provider’s perspective. The primary outcome was the incremental cost-effectiveness ratio (ICER) per quality-adjusted life-year (QALY) gained. One-way and probabilistic sensitivity analyses were performed to assess the robustness of the results.

**Results:** Modelled incidence rates of anal cancer and anogenital warts were significantly reduced following the implementation of 9vHPV vaccine strategies. Vaccinating all MSM aged ≥12 years prevented the most anogenital warts (52.5%) and anal cancer cases (70.4%). Across all MSM vaccination strategies assessed, the ICERs were below the willingness-to-pay (WTP) threshold of USD 50,696 per QALY gained (one-time Hong Kong GDP per capita) or cost-saving, indicating that all 9vHPV strategies were cost-effective. In probabilistic sensitivity analyses, the vaccination strategy for MSM aged ≥12 years consistently demonstrated a high probability of being cost-effective. Furthermore, when the cost of the 9vHPV vaccine decreased or the time horizon was extended, the ICERs for vaccinating MSM aged ≥12 years further declined, making the strategy more cost-effective.

**Conclusion:** Our findings support the implementation of 9vHPV vaccination for MSM in Hong Kong, with vaccinating all MSM aged ≥12 years offering the greatest health and economic benefits.

**Research in context:** *Evidence before this study:* Many countries, such as the United Kingdom, Australia and the United States, have implemented HPV vaccination strategies that include males. In 2018, the UK provided free HPV vaccination for MSM aged ≤45 through sexual health clinics, and later expanded the national HPV programme to include 12–13-year-old boys starting in 2019 to accelerate the reduction of HPV-related cancers in men [1, 2]. This stepwise approach—starting with MSM and later expanding to boys—may be a feasible model for Hong Kong to consider. However, in many settings, the inclusion of males in HPV vaccination programmes is not cost-effective, primarily due to high vaccine coverage among females, which provides indirect protection to males through herd immunity. In Hong Kong, the school-based HPV vaccination program targets only adolescent females. Most cost-effectiveness studies have focused on female-only or gender-neutral strategies without specifically evaluating men who have sex with men (MSM), who are at a substantially higher risk of anal cancer compared to the general population (an estimated 37-fold increased risk in the United States) [3]. HIV-positive MSM are at even greater risk due to their immunocompromised status. We searched PubMed, Google Scholar for articles published between Jan 1, 2000, and March 31, 2024, using terms such as “HPV vaccine”, “MSM”, “cost-effectiveness”, “HIV”, and “Hong Kong”. We found that there is no study evaluating HPV vaccination strategies specifically for MSM in Hong Kong. Existing models primarily address HPV transmission in heterosexual populations. Although a few studies from nearby regions (e.g., mainland China and Singapore) have assessed HPV vaccination among MSM, none have incorporated HIV status in their analyses. These gaps highlight the importance of MSM-specific evaluation of HPV vaccination strategies in Hong Kong.

*Added value of this study:* We did a cost-effectiveness analysis to evaluate 30 vaccine strategies in Hong Kong. These include a combination of 9vHPV at different age groups and HIV status MSM aged ≥12 years. Our findings show that vaccinating all MSM aged ≥12 years is the most cost-effective strategy under base-case assumptions, offering the greatest reductions in the incidence of anogenital warts, anal cancer. This strategy remained the most cost-effective across a range of vaccine costs, particularly when prices decrease and the time horizon increases to 20 years.

*Implications of all the available evidence:* Vaccinating MSM with the 9vHPV is cost-effective in Hong Kong. Among all strategies assessed, vaccinating all MSM aged ≥12 years offers the greatest health and economic benefits, preventing the largest number of HPV-related disease cases while remaining highly cost-effective under the one-time GDP per capita threshold. Although the evaluation from a healthcare provider’s perspective confirms strong cost-effectiveness, affordability remains a key barrier at the individual level—over 50% of MSM consider the high cost of the HPV vaccine a major obstacle to uptake [4]. These findings highlight the importance of publicly subsidized vaccination programs for MSM, similar to how the Hong Kong Childhood Immunisation Programme (HKCIP) provides the 9vHPV vaccine for girls. The findings may also provide valuable policy evidence for other regions or countries that have not yet introduced MSM-specific 9vHPV vaccination strategies.

## Introduction

Human Papillomavirus (HPV) infection is the most common sexually transmitted infection (STI) globally [5]. In men, it commonly appears as anogenital warts, which contribute to considerable health issues and increase the risk of spreading the virus [6]. Compared with the general male population, the prevalence of anal cancers and genital warts was significantly higher in men who have sex with men (MSM) [7]. The high anal cancer incidence among MSM is associated with an elevated anal HPV prevalence, estimated to be 44 times higher than that among the general population [7]. The prevalence of anal cancer is even greater among human immunodeficiency virus positive (HIV-positive) MSM compared to those without HIV [4]. Fortunately, the morbidity and mortality of HPV-related cancers are largely preventable through HPV vaccination, which can significantly reduce morbidity and mortality [8].

Since 2019/20, Hong Kong’s Childhood Immunisation Programme has provided HPV vaccines to eligible female primary school students [9]. However, the incidence of newly diagnosed genital warts among adult men in Hong Kong remains high, at 292.2 cases per 100,000 person-years [10]. Although the incidence of anal cancer (0.4 per 100,000) is relatively low in the general population, MSM has a higher incidence (35 per 100,000 in the US) [11]. Additionally, in the years 2011-2020, MSM accounted for around 83% of HIV cases in Hong Kong. HIV–positive MSM are at particularly high risk of anal HPV infection [7]. These data highlight the burden of HPV-related disease affecting MSM in Hong Kong.

Currently, the 9vHPV vaccine is used in the Childhood Immunisation Programme in Hong Kong. The Gardasil 9 ®: 9-valent HPV (9vHPV) vaccine protects against HPV types 6 and 11, which are associated with the development of genital warts, and HPV types 16, 18, 31, 33, 45, 52, and 58, which are associated with anal cancers [12]. Most high-income countries vaccinate girls aged 9–14 against HPV, but only a few (like Australia, the U.S.) recommend vaccinating boys [13, 14]. When many girls are vaccinated, boys benefit from herd protection, making male vaccination less cost-effective [15]. However, MSM benefit far less from this herd protection [16]. A targeted vaccination program for MSM could help address disparities in disease burden and vaccine access, while remaining potentially cost-effective.

Previous studies have evaluated the cost-effectiveness of female-only and gender-neutral HPV vaccine strategies in Hong Kong [9, 10]. The existing HPV transmission models primarily focus on women or heterosexual men. However, no study has targeted MSM while also incorporating the natural history of anal cancer and HIV status in Hong Kong. In nearby regions, such as mainland China, a cost-effectiveness analysis CEA of HPV vaccination among MSM has been conducted [17], but it did not account for differences in HIV status. Similarly, studies from Singapore, Japan, and South Korea have assessed gender-neutral HPV vaccination strategies, yet none have focused on the MSM population [18–20]. These evaluations find that extending a female vaccination program to males would not be cost-effective in settings with high female vaccine coverage unless female vaccine coverage and/or vaccine prices are sufficiently low. Built on previous findings, this study aimed to assess the cost-effectiveness of implementing HPV vaccination programs for MSM, stratified by different age groups and HIV statuses.

## Methodology

### Study design

We conducted a model-based economic evaluation to determine the cost-effectiveness of 9vHPV MSM HPV vaccination strategies in Hong Kong, adopting a healthcare provider’s perspective. The modelled population included MSM aged ≥12 years residing in Hong Kong. The model was constructed using TreeAge Pro 2019.

### Modelling

A Markov model was constructed to simulate the progression of high-risk HPV infections to anal cancer and subsequent outcomes (death or cure), as well as the course of low-risk infections leading to genital warts or regression to susceptibility (**Fig.1**). The model consisted of 10 health states from susceptible to anal cancer. The simulation was conducted on a cohort of 100,000 MSM (HIV-negative and HIV-positive) stratified into four age groups: aged 12-18, 19-27, 27-44, and ≥45 years. Age groupings were based on differences in sexual activity [21] and the recommended age for HPV vaccination in Hong Kong [22]. The data on MSM age distribution were obtained from a community survey in Hong Kong [23]. The incidence rates of HPV infection were assumed to be different among these four groups (**Supplementary Table S1**). The model projected outcomes over 10 years. The main parameters used to inform this model are shown in **Table 1**.

**Fig. 1.**
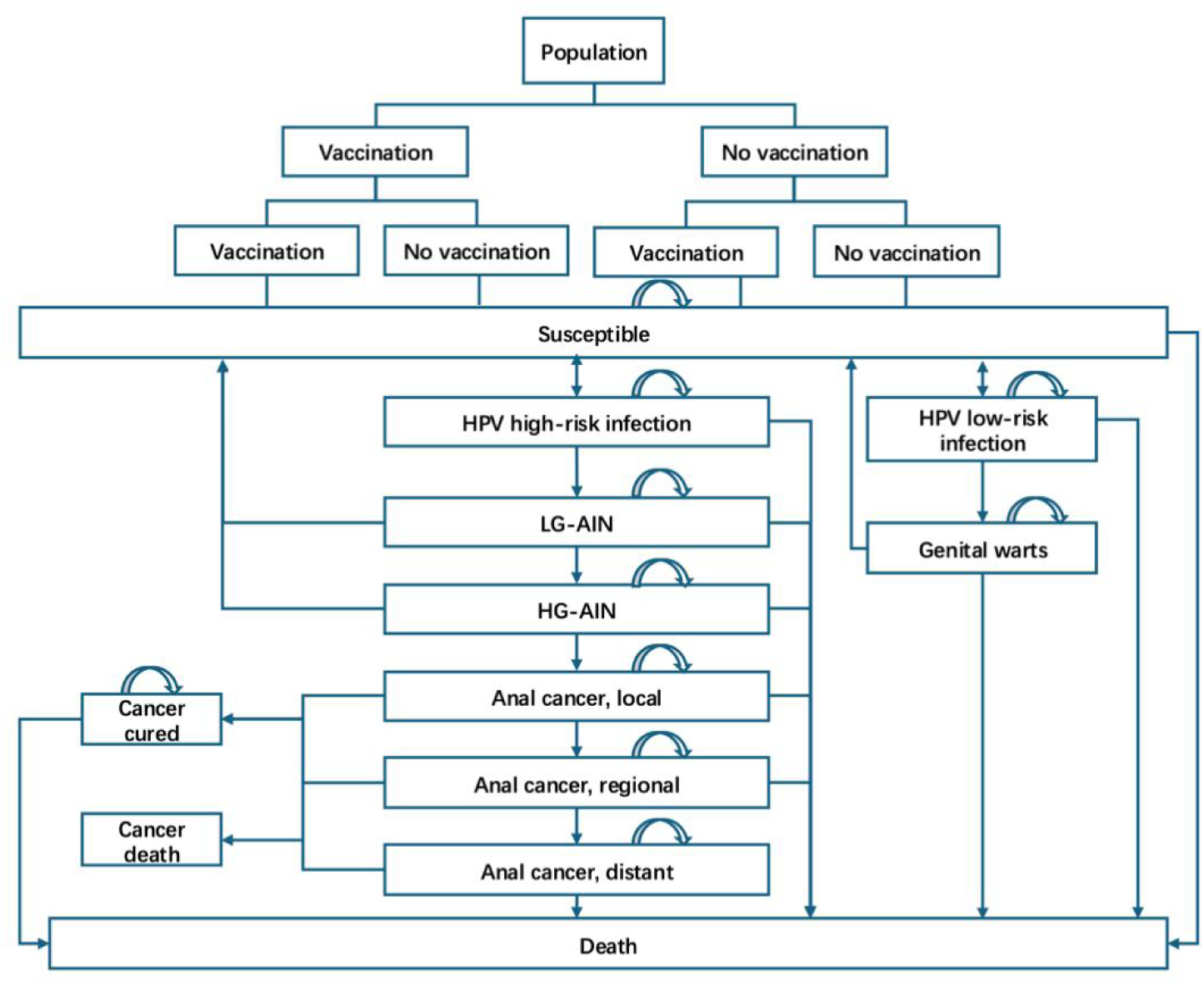
Markov model structure. HIV: Human Immunodeficiency Virus; HPV: Human Papillomavirus; LG-AIN: Low-Grade Anal Intraepithelial Neoplasia; HG-AIN: High-Grade Anal Intraepithelial Neoplasia.

**Table 1.**
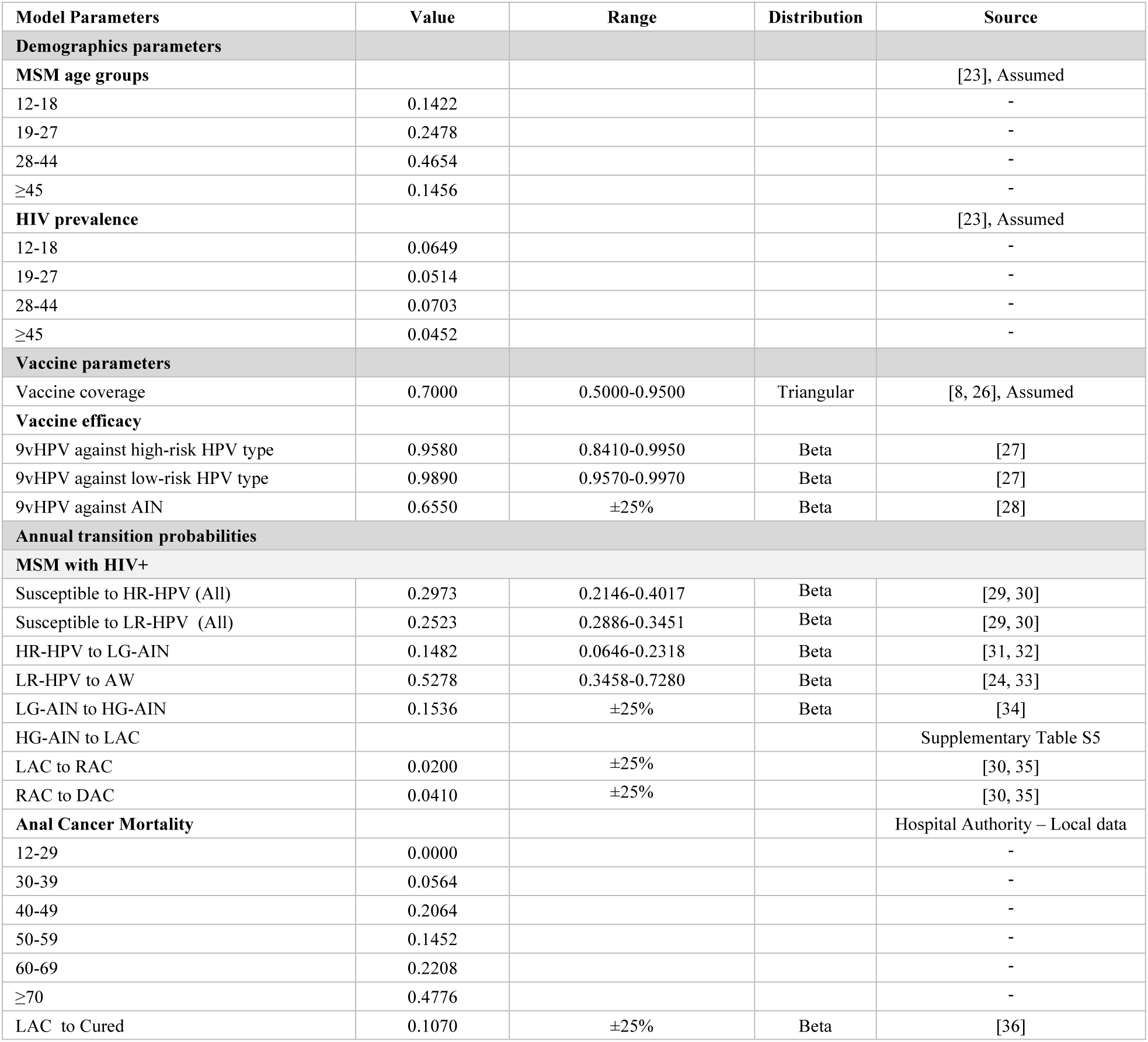
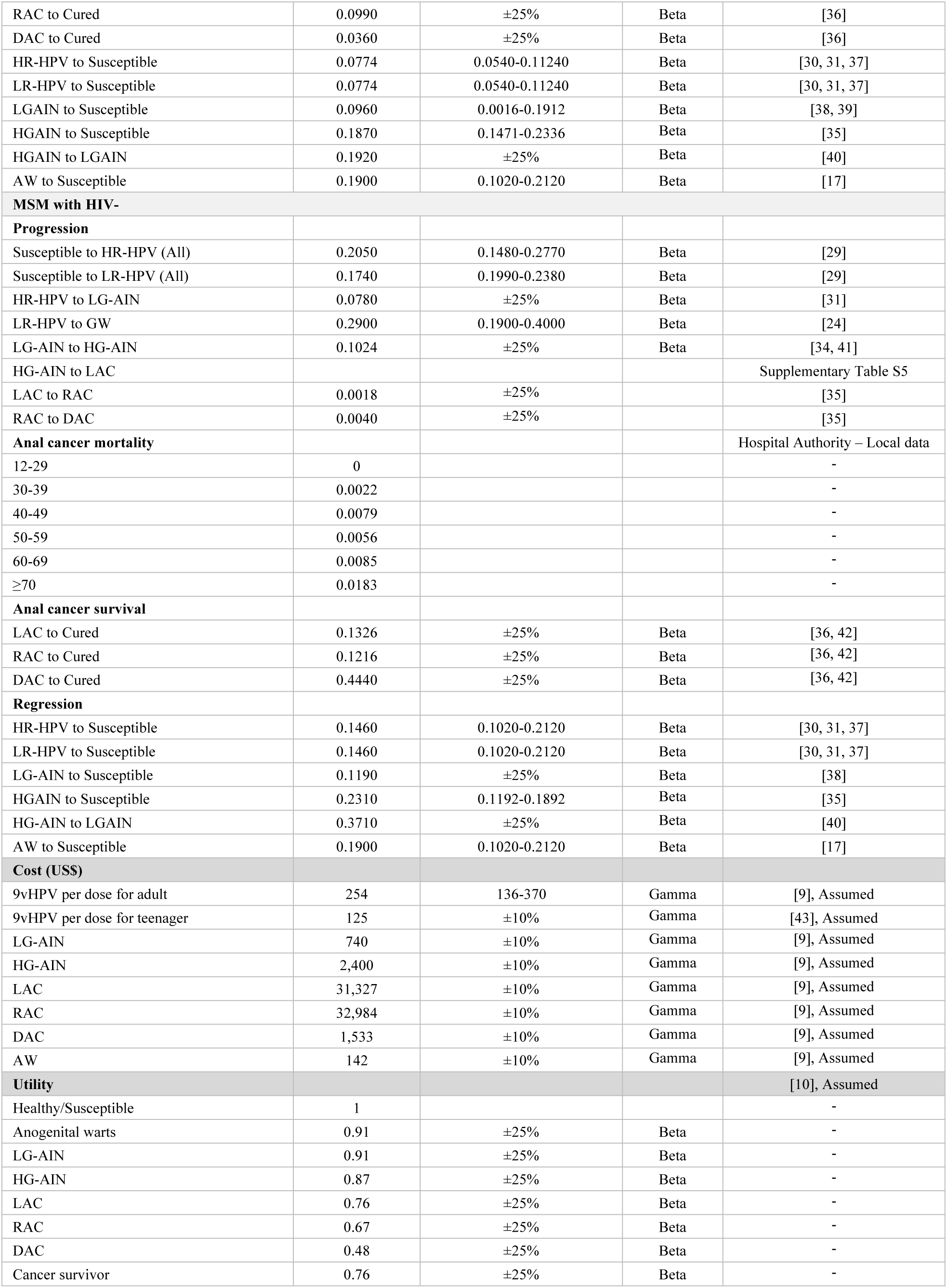
Markov model inputs. HIV: Human Immunodeficiency Virus; HPV: Human papillomavirus; MSM: Men who have sex with men; HR-HPV: High-risk human papillomavirus; LR-HPV: Low-risk human papillomavirus; LG-AIN: Low-grade anal intraepithelial neoplasia; HG-AIN: High-grade anal intraepithelial neoplasia; AW: anogenital warts; 9vHPV: 9-valent human papillomavirus vaccine

Data used in the model were sourced from official government websites, the Hospital Authority, and published literature. Details about all parameters applied in the model are presented in **Table 1**. For the sensitivity analysis, the range of values tested for each parameter was based either derived from the 95% confidence intervals reported in the referenced studies or estimated by applying a ±25% variation from the base-case value for the transition probabilities [24] and a ±10% variation for cost parameters [9]

Individuals entered the model from a ‘susceptible’ state. Each susceptible individual could be infected with either low-risk or high-risk HPV subtypes [17], remain in their current health state, or die. For MSM, infection with a high-risk HPV type could lead to the development of LG-AIN. Individuals with LG-AIN could either progress to HG-AIN or regress to the susceptible state. Further progression of HG-AIN could lead to anal cancer, or these conditions could regress to the LG-AIN or susceptible state. The anal cancer stage consisted of local, regional, and distant cancer states. Age-specific natural background mortality rates were obtained from a previous local study in Hong Kong (**Supplementary Table S2**) [9]. Individuals diagnosed with cancer were subject to both age-specific cancer-related mortality (**Table 1**) and the natural background death rate. The model operated on a 1-year cycle length, with a half-cycle correction applied.

The different age groups of MSM enter the model simultaneously to better fit the real-world epidemic situation of HPV-related diseases among MSM in Hong Kong. We assumed that 14.2%, 24.8%, 46.5%, and 14.6% of MSM were in age groups 12-18, 19-27, 28-44, and ≥45 respectively [25], from the beginning of our model simulation, and our model assumes the proportion keeps constant **(Supplementary Table S3).**

### Intervention strategies

We defined the 23% coverage of 9vHPV vaccination for MSM as the status quo in Hong Kong [4], given that the current Hong Kong HPV vaccination program targets only school-aged girls, leaving boys unvaccinated [9]. The HPV vaccine is accessible to MSM through private clinics [4].

30 intervention strategies (**Supplementary Table S4**) were investigated in our study. These included various 9vHPV vaccination strategies targeting MSM, stratified by age group (12-18, 19–27, 28-45, and >45 years) and HIV status (positive and negative). We assumed the two-dose 9vHPV for MSM aged 12–14 years would be Hong Kong Childhood Immunisation Programme (HKCIP)-based and 14-18 would be catch-up program based. The 9vHPV vaccine coverage of MSM aged 12-18 is assumed to reach 70% [8–10]. HIV-positive MSM aged 12-18 years have to receive three doses of vaccination [44]. For MSM aged over 18, a three-dose regimen was considered, with the same assumed coverage rate of 70% [8, 9, 45]. In our model, vaccination was assumed to occur at the time the cohort entered the model. The model assumed that HPV vaccines provide 10-year protection [46], and a two-dose 9vHPV vaccination schedule is non-inferior to a three-dose schedule [47, 48]

### Data analysis

The HPV infection incidence of high-risk HPV infection and low-risk HPV infection is different for age groups and HIV status among MSM. The probabilities of annual transition were derived from published literature on the natural history of anal cancer and anogenital warts (**Table 1**). Anal cancer mortality rate (**Table 1**) and the age-specific all-cause mortality rate (**Supplementary Table S2**) were obtained from the Hong Kong Cancer Registry, Hospital Authority.

The costs of the vaccine program included the costs of the vaccine, diagnosis, and treatment (**Table 1**). The cost data in this study were primarily obtained from previous publications and the Hospital Authorities’ records. The model utilized FPAHK (a non-profit organization) pricing for the HPV vaccines, with per-dose costs of US$125 for 9vHPV for 12-18 MSM (adolescent) [49], and assumes average private clinics pricing in US$254 for >18 MSM (adult) [50]. All costs are from a healthcare payer perspective. Costs were converted from Hong Kong dollars to US dollars (US$ 1 = HKD 7.829, in 2023) [51].

Utility scores for HPV-related states were obtained from a cost-effectiveness analysis in Hong Kong (**Table 1**) [10], which included the general population’s health status and HPV-related disease status. We assumed a discount rate of 3% for both (quality-adjusted life years) QALYs and cost. As HPV infections are generally asymptomatic, we assumed HPV high-risk infection and low-risk infection states had the same utility weight as being in the susceptible state [17]. Due to the limited avalibility of utility weights of LG-AIN and HG-AIN, we assumed utility weights to be the same as cervical intraepithelial neoplasia (CIN) in Hong Kong (**Table 1**) [16].

We compared each MSM vaccination strategy to the no-routine MSM vaccination strategy by calculating their incremental costs and QALYs. From these comparisons, we established the cost-effectiveness frontier and determined the ICER,. This analysis helped identify the most cost-effective MSM vaccination strategy. Cost-effectiveness was assessed using WHO criteria, categorizing strategies as highly cost-effective, cost-effective, or not cost-effective based on ICER thresholds of less than 1, between 1 and 3, or greater than 3 times the GDP per capita. In 2023, Hong Kong’s GDP per capita was US$ 50,696 [52].

### Sensitivity analyses

We conducted one-way sensitivity analyses to determine how the ICER was affected by varying the following input parameters: utility for HPV-associated disease (±25%), vaccine price (±10%), disease treatment costs (±10%), duration of vaccine-mediated protection (set to 10 years), and transition probability (±25% or confidence interval range), to understand how the ICER depends on uncertainty in the model parameters [9, 24]. Each analysis was conducted independently, with all other variables set to their base-case values. In probabilistic sensitivity analyses, we simultaneously varied multiple key input parameters across prespecified distributions over 1,000 simulations. For all costs, a Gamma distribution was used, and for all transition probabilities and utilities, a Beta distribution (0–1) was used [53].

### Scenario analysis

The cost of the 9vHPV vaccine is a critical factor in cost-effectiveness studies [54]. We examined the potential effects of both increasing and decreasing the cost of the 9vHPV vaccine in our base-case analyses. This involved (i) assuming price changes per dose for MSM (both adolescent and adult) ± 25% and ± 50%. We also considered the following alternative scenarios: (ii) Vaccine protection wanes completely after a fixed duration of 10 years; (iii) the time horizon over which vaccination costs and benefits were assumed to accrue (5 years, 20 years);

## Results

### Base-case analyses

Modelled incidence rates of anal cancer and anogenital warts were significantly reduced following the implementation of 9vHPV vaccine strategies. The strategies that prevented the most cases of HPV-related diseases were identified across all different age-stratified HPV vaccination strategies. Among one-group-targeted strategies, vaccinating MSM aged 28–44 years prevented the most anogenital warts (26.7%). The two-group-targeted strategy vaccinating MSM aged 19–27 & 28–44 years prevent the most cases, 38.8% of anogenital warts and 58.0% of anal cancer. The three-group-targeted strategy covering MSM aged 12– 44 years yielded the greatest reduction in anogenital warts (45.8%). Expanding vaccination coverage across more age groups resulted in greater reductions in HPV-related diseases cases. Vaccinating all MSM ≥ 12 prevented about 75.6 million cases of anogenital warts (52.5% reduction) and 143,000 anal cancer cases (70.4% reduction) (**Supplementary Table S8**).

We analyzed the incremental costs and incremental QALYs for 30 vaccination strategies targeting MSM, compared to a no-routine 9vHPV scenario, in a cohort of 100,000 MSM over a 10-year horizon (**Table 2**). With a willingness-to-pay threshold at one-time the Hong Kong per-capita GDP (US$ 50,696 per QALY gained), all 9vHPV vaccination strategies were highly cost-effective. The targeted vaccination strategies among HIV-positive individuals across different age groups would result in cost savings ranging from US$ 720,000 to US$ 4,520,000 and result in 500–3200 QALYs gained. The most cost-saving strategy woule be vaccination for HIV-positive MSM aged 12-18. Broader strategies targeting all MSM in certain age groups would incur an incremental cost of US$ 820,000 to 215,950,000, with QALY gained from 3,700 to 30,500. The strategy of vaccination for all MSM aged ≥12 years would increase the QALYs most (30,500 QALYs).

**Table 2.** ICERs (HKD per QALY gained) for base-case analyses. . ICER: incremental cost-effectiveness ratio; HKD: Hong Kong dollar; QALY: quality-adjusted life year. Cost-saving (costs less and is more effective than base case strategy)

| Strategy |  | Cost<br>(*10<sup>6</sup>) | Incremental<br>cost (*10<sup>6</sup>) | QALYs (*10<sup>6</sup>) | Incremental<br>QALYs<br>(*10<sup>6</sup>) | ICER |
| --- | --- | --- | --- | --- | --- | --- |
| No-routine vaccine |  | 349.44 |  | 7.8307 |  | -367.57 |
| Population | Age groups |  |  |  |  |  |
| HIV+ | 12-18 | 344.92 | -4.52 | 7.8312 | 0.0005 | Cost saving |
|  | 12-18 & ≥45 | 345.61 | -3.83 | 7.8315 | 0.0008 | Cost saving |
|  | 12-18 & 28-44 | 345.98 | -3.46 | 7.8334 | 0.0027 | Cost saving |
|  | ≥45 | 346.25 | -3.19 | 7.8311 | 0.0003 | Cost saving |
|  | 12-27 | 346.32 | -3.12 | 7.8319 | 0.0011 | Cost saving |
|  | 28-44 | 346.62 | -2.82 | 7.8329 | 0.0022 | Cost saving |
|  | 12-18 & ≥28 | 346.67 | -2.77 | 7.8337 | 0.0030 | Cost saving |
|  | 19-27 | 346.96 | -2.48 | 7.8314 | 0.0007 | Cost saving |
|  | 12-27 & ≥45 | 347.02 | -2.42 | 7.8322 | 0.0015 | Cost saving |
|  | ≥28 | 347.31 | -2.13 | 7.8333 | 0.0025 | Cost saving |
|  | 12-44 | 347.39 | -2.05 | 7.8341 | 0.0034 | Cost saving |
|  | 19-27 & ≥45 | 347.66 | -1.78 | 7.8317 | 0.0010 | Cost saving |
|  | 19-44 | 348.03 | -1.41 | 7.8336 | 0.0029 | Cost saving |
|  | ≥12 | 348.08 | -1.36 | 7.8344 | 0.0037 | Cost saving |
|  | ≥19 | 348.72 | -0.72 | 7.8339 | 0.0032 | Cost saving |
| All | 12-18 | 350.26 | 0.82 | 7.8344 | 0.0037 | 224.84 |
|  | ≥45 | 384.87 | 35.43 | 7.8343 | 0.0036 | 9838.81 |
|  | 12-18 & ≥45 | 389.58 | 40.14 | 7.8380 | 0.0073 | 5527.64 |
|  | 19-27 | 412.56 | 63.12 | 7.8370 | 0.0063 | 10056.74 |
|  | 12-27 | 417.26 | 67.82 | 7.8407 | 0.0099 | 6825.70 |
|  | 19-27 & ≥45 | 451.87 | 102.43 | 7.8406 | 0.0099 | 10370.50 |
|  | ≥28-44 | 452.36 | 102.92 | 7.8476 | 0.0169 | 6085.30 |
|  | 12-27 & ≥45 | 456.58 | 107.14 | 7.8443 | 0.0135 | 7914.18 |
|  | 12-18 & 28-44 | 457.07 | 107.63 | 7.8513 | 0.0206 | 5231.39 |
|  | ≥28 | 491.68 | 142.24 | 7.8512 | 0.0205 | 6933.59 |
|  | 12-18 & ≥28 | 496.39 | 146.95 | 7.8549 | 0.0242 | 6078.45 |
|  | 19-44 | 519.36 | 169.92 | 7.8539 | 0.0232 | 7327.65 |
|  | 12-44 | 524.07 | 174.63 | 7.8576 | 0.0268 | 6503.97 |
|  | ≥19 | 558.68 | 209.24 | 7.8575 | 0.0268 | 7810.21 |
|  | ≥12 | 563.39 | 213.95 | 7.8612 | 0.0305 | 7025.94 |

### Sensitivity analyses

The tornado diagram in **Fig. 2** illustrates the results of our one-way sensitivity analysis comparing the cost-effectiveness of the vaccination strategy targeting all MSM aged ≥12 years against the no-routine vaccination for MSM. Across the tested sensitivity ranges, none of the parameters caused the ICER to exceed US$ 50,696 per quality-adjusted life year (QALY) gained. Among all variables, the cost of 9vHPV vaccine had the greatest impact on ICER. Other variables were transition probabilities, disease utility, efficacy of 9vHPV against HPV high risk type.

**Fig. 2:**
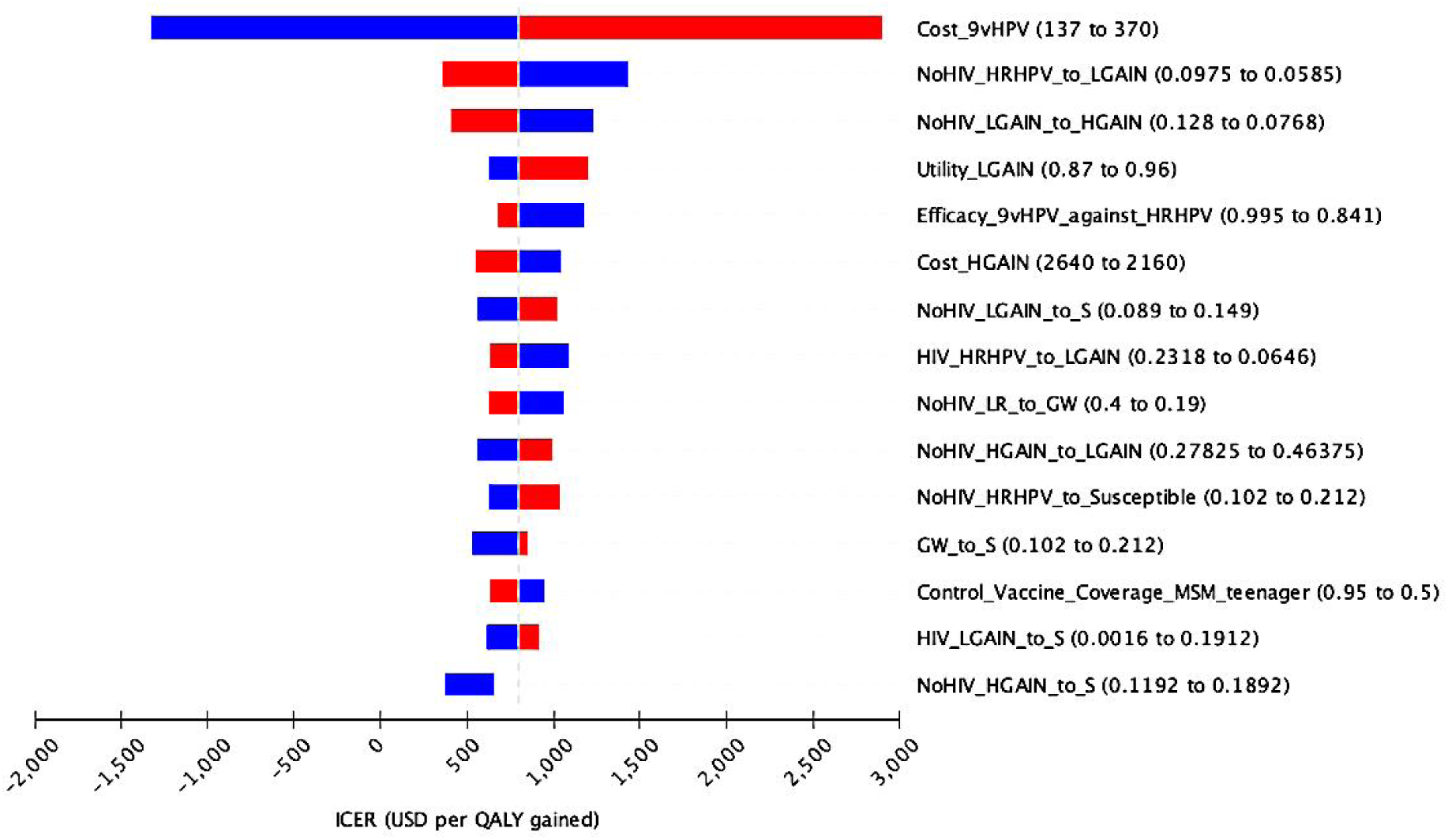
Tornado diagram for one-way sensitivity analyses of key model parameters. Blue/red bars are indicative of sensitivity values below/above the base case. The strategy of vaccinating all MSM aged ≥12 years against the no routine vaccination scenario. 9vHPV, 9-valent human papillomavirus vaccine; HRHPV, high-risk HPV; LRHPV, low-risk HPV; ICER, incremental cost-effectiveness ratio; QALY, quality-adjusted life year; MSM, men who have sex with men; HIV, human immunodeficiency virus; HGAIN, high-grade anal intraepithelial neoplasia; LGAIN, low-grade anal intraepithelial neoplasia; S, susceptibility.

**Fig. 3** shows the cost-effectiveness acceptability curves for all vaccination strategies targeting MSM WTP thresholds ranging from US$ 0 to US$ 50,696 per QALY gained (one-time the per-capita GDP). At a WTP of US$ 0 per QALY, the most acceptable strategy was vaccinating HIV-positive MSM aged 12–18 & 28–44 years (50.4%), followed by HIV-positive MSM aged 12–18 only (42.5%). As WTP increased to US$ 5,069.6, targeting all HIV-positive MSM aged ≥12 years became most acceptable (50.3%). At US$ 10,139.2, the strategy targeting all MSM aged ≥12 years dominated (65.5%). From US$15,208.8 onwards, this strategy consistently remained the most acceptable, reaching full acceptability (100%) at WTP ≥ US$ 20,278.4 per QALY.

**Fig 3.**
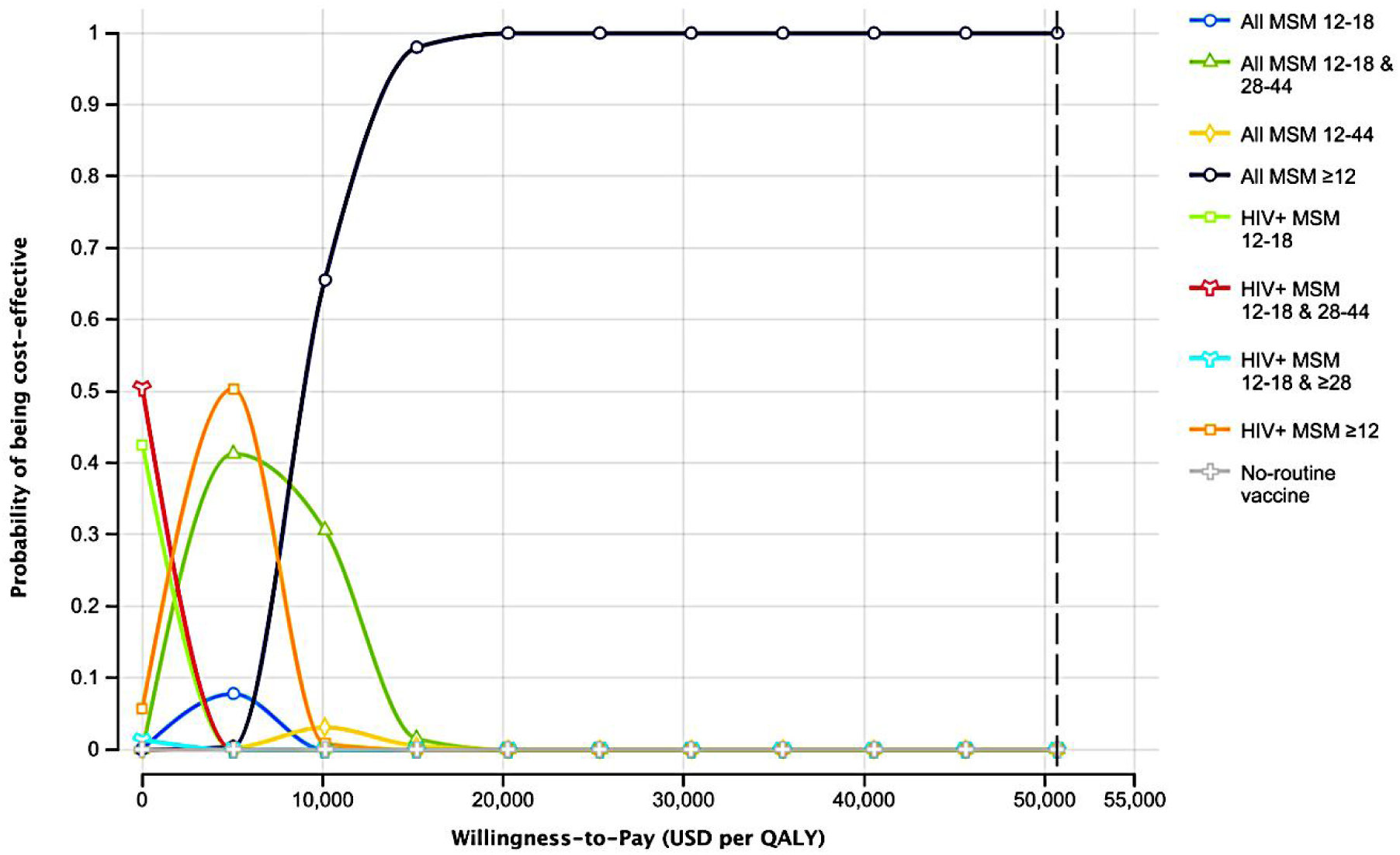
Cost-effectiveness acceptability curves for all strategies. Intervention strategies that never have the probability of being cost-effective within the willingness-to-pay threshold of one-time the Hong Kong per-capita GDP are not shown. QALYs: quality-adjusted life-years; GDP: gross domestic product.

We evaluated how different time horizons, under a 10-year vaccine efficacy assumption, affect the most cost-effective strategies among MSM (**Fig. 4**). When the time horizon was shortened to 5 years, the most acceptable strategy at low WTP levels was targeting HIV-positive MSM aged 12–18 (100%). As WTP increased to US$ 10,139.2 and above, broader strategies—vaccinating HIV-positive MSM aged 12–18 & 28–44 or HIV-positive MSM aged ≥12—became more acceptable. At the WTP equal to one-time the per-capita GDP, the strategy of 9vHPV for all MSM aged ≥12 became increasingly acceptable (rising to 55.9% at US$ 50,696). In contrast, when the time horizon was extended to 20 years, the strategy targeting all MSM aged ≥12 years quickly became dominant. From a WTP of US$ 5,069.6 per QALY onwards, this strategy had high probability of being cost-effective, reaching 100% at US$ 10,139.2 and remaining dominant across all higher WTP levels.

**Fig. 4.**
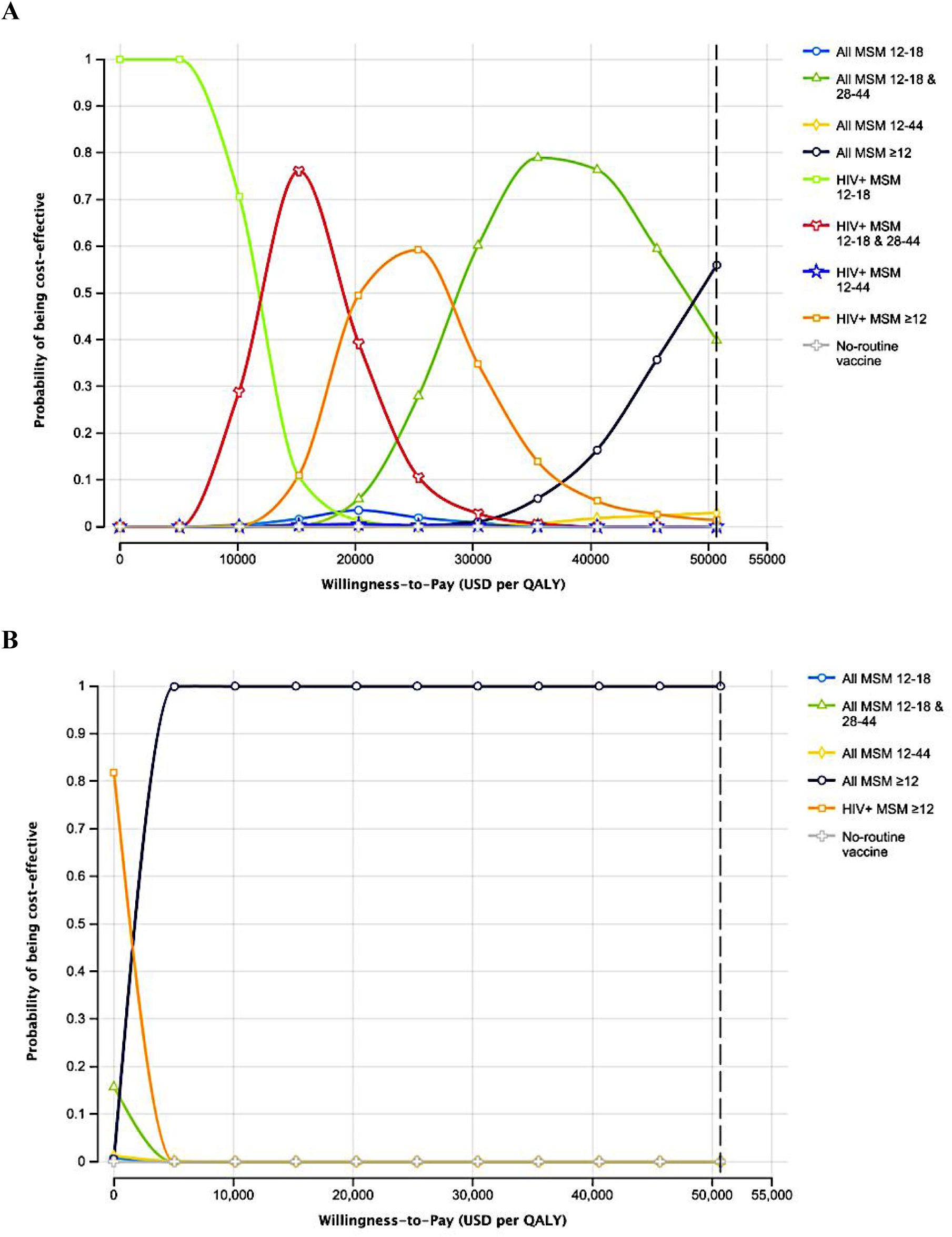
Cost-effectiveness acceptability curves for all strategies under different time horizons. A: 5-year time horizon; B: 20-year time horizon. Each figure shows the probability of each intervention strategy being the cost-effective across US$ 0-50,696 WTP thresholds, assuming a 10-year vaccine efficacy. Intervention strategies with negligible probability of being cost-effective across all WTP thresholds are not shown. QALYs: quality-adjusted life-years.

## Discussion

The Hong Kong Childhood Immunisation Programme has made significant progress. In the school years 2023/24, the two-dose coverage rates of HPV vaccination for Primary Six school girls reached 91% [44]. As female coverage rates increase, it becomes more challenging to demonstrate the additional benefits of vaccinating males [10]. Although a cost-effectiveness study in Hong Kong has demonstrated that a gender-neutral 9vHPV vaccination strategy is cost-effective, it did not specifically account for MSM [10]. This may lead to an overestimation of the overall cost-effectiveness of the gender-neutral strategy, as MSM bear a disproportionately higher burden of HPV-related diseases [55] and do not benefit from female herd immunity [56]—even when female vaccination coverage is high. To address this gap, our analysis assessed the health and economic outcomes of implementing a 9vHPV vaccination programme for MSM in Hong Kong. Among all strategies evaluated, vaccinating all MSM aged ≥12 years emerged as the most cost-effective option. Moreover, reduction in vaccine cost or extending the time horizon would further improve the cost-effectiveness of this strategy.

The UK’s national HPV vaccination programme has already offers MSM aged up to 45 the vaccine through Specialist Sexual Health Services and HIV clinics [57]. Our findings are consistent with the studies on the cost-effectiveness of the quadrivalent HPV vaccine among MSM (with and without HIV) in the UK [16]. We found that broader targeting of MSM becomes more cost-effective when vaccine costs are lower. Conversely, when vaccine costs are doubled in the UK, targeting HIV-positive MSM—yields better value. However, our sensitivity analysis assuming a ±50% variation in vaccine cost (**Supplementary Table S9**) did not significantly alter the results. This likely reflects differences in cost of HPV-related diseases. The UK study assumed much lower costs for treating anal cancer. In contrast, the treatment cost for anal cancer in Hong Kong’s public hospitals is nearly four times higher than in the UK. This cost difference enhances the economic value of vaccination. For instance, if the cost of treating each case of anal cancer is very high, even if the vaccine itself is relatively expensive, the long-term healthcare savings from preventing cancer can offset the initial costs of vaccination. Additionally, we assumed a higher vaccine efficacy for the HPV vaccine in our model. Since the HKCIP in Hong Kong provides the 9vHPV vaccine to girls and has discontinued the supply of the 2vHPV and 4vHPV vaccines due to low demand, we used 9vHPV for MSM in our analysis. The 9vHPV vaccine offers protection against more high-risk HPV subtypes, making it more effective in preventing anal cancer [58]. This contributed to more favorable cost-effectiveness outcomes.

We distinguished between the vaccine cost for adolescents and adults, as the Hong Kong Childhood Immunisation Programme and its catch-up programme currently provide free-of-charge HPV vaccination to individuals up to 18 years. Therefore, MSM aged ≤18 are assumed to receive the 9vHPV vaccine through the public system, while MSM aged >18 are assumed to receive 9vHPV vaccine through private clinics, resulting in substantially higher costs for adults. However, according to a local survey on MSM in Hong Kong [23] the proportion of MSM aged 12–18, and the prevalence HIV among MSM aged 12–18 is relatively low. In addition, previous studies—such as a HPV epidemiology study among MSM in China [17] and a U.S.-based cost-effectiveness analysis of 9-valent HPV vaccination [21] have found that MSM aged 28–44 exhibit the highest rates of HPV infection. As such, focusing vaccination efforts solely on MSM aged 12–18 may not be the most cost-effective strategy, despite its potential probability of being cost-effective under very low WTP thresholds.

We used 10-year time horizon, because the clinical trials showed that the HPV vaccine efficacy is up to 10 years [59]. In our sensitivity analysis, we varied the time horizon to 5 years and 20 years while keeping the vaccine protection duration fixed at 10 years. When a 5-year time horizon was used, vaccinating all MSM aged ≥12 years became less cost-effective compared to the base case analysis. This is because the costs of HPV vaccination are incurred immediately, but the full benefits of vaccination—such as the prevention of HPV-related diseases—take many years to materialize. As a result, shorter time horizons make vaccination appear less favorable in terms of cost-effectiveness [60]. In contrast, with a 20-year time horizon, vaccinating all MSM aged ≥12 years became more cost-effective and dominant. Despite the vaccine’s protection waning completely after 10 years, the long-term benefits of reduced HPV-related disease incidence still made vaccination for all MSM a cost-effective strategy over the extended period.

An important limitation of this study was the lack of local data for several key parameters, including transition probabilities, health utility values, and the most recent age distribution and HIV prevalence among MSM in Hong Kong. As a result, we had to make several assumptions based on meta-analyses, regional data from nearby areas such as Shenzhen, or extrapolations from international studies. Although our ICER estimates were not highly sensitive to variations in these parameters, the use of stronger empirical data would enhance the robustness and credibility of our findings. In particular, availability of age- and risk-stratified data on sexual activity and HPV prevalence among MSM would facilitate more granular modelling of 9vHPV vaccination strategies. It is likely that the vaccine would be even more cost-effective in subgroups with higher sexual risk, such as highly sexually active MSM.

All 9vHPV vaccination strategies targeting MSM living with and without HIV are cost-effective means of preventing anal cancer under Hong Kong’s current per capita GDP. However, a 2025 study on HPV vaccine uptake and service preferences among MSM in Hong Kong highlighted that the primary barrier to vaccination was the high out-of-pocket cost. The study suggested that reducing the cost to below USD 128 per dose could significantly improve vaccine uptake among MSM. Notably, this price aligns with the current cost of HPV vaccination for girls aged 12–18 under the female-only programme. This raises important policy and ethical considerations regarding the integration of MSM into publicly funded HPV vaccination programmes.

Questions of fairness and equity emerge—particularly in determining whether male adolescents aged 12–18 who identify as or may become MSM should be included, and how to do so without reinforcing social stigma or discrimination. Identifying MSM status at a young age is challenging due to both developmental and social factors, and the risk of stigma may deter individuals from accessing vaccination services. Indeed, it may be argued that requesting 12-year-olds to identify or define their own sexuality at an age when they are not fully developed could be viewed as potentially socially coercive and aggravate anxieties and mental health challenges. A local survey [4] found that most MSM preferred receiving HPV vaccination at private clinics or NGOs, suggesting a strong preference for privacy and inclusive, gay-friendly healthcare settings. This underscores the importance of reducing stigma and enhancing service accessibility by fostering a more welcoming environment and improving the quality of vaccination services in private sectors. Evidence from Australia and the UK shows that MSM responded well to free HPV vaccination programs. Hong Kong could consider similar initiatives to boost vaccine uptake in this grou [61]. Following the UK model, offering HPV vaccination to MSM attending sexual health or HIV testing services may further enhance coverage. Policy strategies aimed at reducing cost and stigma and expanding access to HPV vaccination across at-risk male populations, are the priorities.

## Conclusion

Our findings support the implementation of 9vHPV vaccination for MSM in Hong Kong, with vaccinating all MSM aged ≥12 years offering the greatest health and economic benefits.

## Data Availability

The original contributions presented in this study are included in the article/supplementary material. Further inquiries can be directed to the corresponding author(s).

## Acknowledgements

None.

