## Supplementary appendix for "Evaluating Cost-effectiveness of 9-valent HPV Vaccination for Men Who Have Sex with Men by HIV Status in Hong Kong"

**lementary Table S1. Annual HPV infection rates by age group and HIV status in MSM.**

| Age Group | HIV+/HR-HPV | HIV+/LR-HPV | HIV−/HR-HPV | HIV−/LR-HPV |
|---|---|---|---|---|
| 12–18 | **0.2027** | **0.1720** | **0.1398** | **0.1186** |
| 19–27 | **0.2027** | **0.1720** | **0.1398** | **0.1186** |
| 28–44 | **0.4054** | **0.3440** | **0.2796** | **0.2372** |
| ≥45 | **0.2027** | **0.1720** | **0.1398** | **0.1186** |

MSM = men who have sex with men; HPV = human papillomavirus; HR-HPV = high-risk human papillomavirus; LR-HPV = low-risk human papillomavirus; HIV = human immunodeficiency virus.

The overall HPV incidence is from a an observational cohort study [1]. To derive age-specific infection rates for high risk HPV types and low risk HPV types and HIV statuses among MSM, we used the following formula based on age group weighting (Supplementary Table S3) and relative incidence: $P_i = k \times r_i$ (k=Total susceptibility $/\sum w_i \times r_i$ )

To estimate the incidence of high-risk (HR) and low-risk (LR) HPV in HIV-positive MSM, we used the prevalence ratio between HIV-positive (92.6%) and HIV-negative (63.9%) MSM [2], giving a ratio of approximately 1.45. Assuming susceptibility increases proportionally with prevalence, we multiplied the known susceptibility in HIV-negative MSM (HR-HPV: 0.0171; LR-HPV: 0.0142) by this ratio. The estimated susceptibilities in HIV-positive MSM are therefore 0.0248 for HR-HPV and 0.0206 for LR-HPV. This simplified approach assumes a direct relationship and may not account for other influencing factors.

The incidence rates of HPV infection were assumed to be different among these four groups. The age-stratified HPV incidences were assumed to be 1:1:2:1 among the 12-18, 19-27, 28-44 and ≥45 age groups, to reflect the higher overall incidences of genital warts and anal cancer among the intermediate male age group [3].

**Supplementary Table S2. Age-specific natural background mortality rates.**

| Annual all-cause mortality rate by HIV status and age (%). | HIV- | HIV+ |
|---|---|---|
| 11–14 years | 0.0083 | 0.018094 |
| 15–19 years | 0.0215 | 0.04687 |
| 20–24 years | 0.0322 | 0.070196 |
| 25–29 years | 0.0466 | 0.101588 |
| 30–34 years | 0.0622 | 0.135596 |
| 35–39 years | 0.0833 | 0.181594 |
| 40–44 years | 0.1253 | 0.273154 |
| 45–49 years | 0.1990 | 0.4338 |
| 50–54 years | 0.3107 | 0.677326 |
| 55–59 years | 0.5293 | 1.153874 |
| 60–64 years | 0.8374 | 1.825532 |
| 65–69 years | 1.2641 | 2.755738 |
| 70–74 years | 2.1524 | 4.692232 |
| 74–79 years | 3.4555 | 7.53299 |
| 80–84 years | 6.0256 | 13.135808 |
| 85+ years | 12.2915 | 26.79547 |

HIV = human immunodeficiency virus; HIV− = HIV-negative individuals; HIV+ = HIV-positive individuals

The monthly all-cause mortality rates for HIV-negative MSM were based on age-specific mortality rates for males in Hong Kong [4]. For HIV-positive MSM, a higher mortality risk was applied by incorporating a monthly mortality risk ratio of 2.18, as reported in a previous study [5]

**Supplementary Table S3. The proportion of MSM age groups**

| Age Group | Proportion |
|---|---|
| 12–18 | **0.1422** |
| 19–27 | **0.2478** |
| 28–44 | **0.4654** |
| ≥45 | **0.1456** |

To adjust the proportions according to the newly defined age groups (12-18, 19-27, 28-44, ≥41), we'll need to redistribute the existing age group data (≤20, 21-30, 31-40, ≥41) into the new categories. We'll assume a uniform distribution within each group for this calculation [6].

| MSM age groups | [7] |
|---|---|
| 12-20 | **9.6%** |
| 21-30 | **42%** |
| 31-40 | **32.9%** |
| ≥41 | **15.6%** |

**Supplementary Table S4. Age-specific natural background mortality rates.**

| Annual all-cause mortality rate by HIV status and age (%). | HIV- | HIV+ |
|---|---|---|
| 11–14 years | **0.0083** | **0.018094** |
| 15–19 years | **0.0215** | **0.04687** |
| 20–24 years | **0.0322** | **0.070196** |
| 25–29 years | **0.0466** | **0.101588** |
| 30–34 years | **0.0622** | **0.135596** |
| 35–39 years | **0.0833** | **0.181594** |
| 40–44 years | **0.1253** | **0.273154** |
| 45–49 years | **0.1990** | **0.4338** |
| 50–54 years | **0.3107** | **0.677326** |
| 55–59 years | **0.5293** | **1.153874** |
| 60–64 years | **0.8374** | **1.825532** |
| 65–69 years | **1.2641** | **2.755738** |
| 70–74 years | **2.1524** | **4.692232** |
| 74–79 years | **3.4555** | **7.53299** |
| 80–84 years | **6.0256** | **13.135808** |
| 85+ years | **12.2915** | **26.79547** |

HIV = human immunodeficiency virus; HIV− = HIV-negative individuals; HIV+ = HIV-positive individuals

The monthly all-cause mortality rates for HIV-negative MSM were based on age-specific mortality rates for males in Hong Kong [4]. For HIV-positive MSM, a higher mortality risk was applied by incorporating a monthly mortality risk ratio of 2.18, as reported in a previous study [5]

**Supplementary Table S4. HPV vaccination strategies for MSM.**

| Strategy | Age groups | | | |
|---|---|---|---|---|
| | **12-14** | **15-27** | **28-44** | **≥45** |
| One-group-targeted strategies | | | | |
| All MSM | **1** | **0** | **0** | **0** |
| | **0** | **1** | **0** | **0** |
| | **0** | **0** | **1** | **0** |
| | **0** | **0** | **0** | **1** |
| HIV+ MSM | **1** | **0** | **0** | **0** |
| | **0** | **1** | **0** | **0** |
| | **0** | **0** | **1** | **0** |
| | **0** | **0** | **0** | **1** |
| Two-group-targeted strategies | | | | |
| All MSM | **1** | **1** | **0** | **0** |
| | **1** | **0** | **1** | **0** |
| | **1** | **0** | **0** | **1** |
| | **0** | **1** | **1** | **0** |
| | **0** | **1** | **0** | **1** |
| | **0** | **0** | **1** | **1** |
| HIV+ MSM | **1** | **1** | **0** | **0** |
| | **1** | **0** | **1** | **0** |
| | **1** | **0** | **0** | **1** |
| | **0** | **1** | **1** | **0** |
| | **0** | **1** | **0** | **1** |
| | **0** | **0** | **1** | **1** |
| Three-group-targeted strategies | | | | |
| All MSM | **1** | **1** | **1** | **0** |
| | **1** | **1** | **0** | **1** |
| | **1** | **0** | **1** | **1** |
| | **0** | **1** | **1** | **1** |
| HIV+ MSM | **1** | **1** | **1** | **0** |
| | **1** | **1** | **0** | **1** |

| | | | | |
|---|---|---|---|---|
| | **1** | **0** | **1** | **1** |
| | **0** | **1** | **1** | **1** |
| Three-group-targeted strategies | | | | |
| All MSM | **1** | **1** | **1** | **1** |
| HIV+ MSM | **1** | **1** | **1** | **1** |

MSM = men who have sex with men; Number "0" represents no vaccine, "1" represents vaccine.

**Supplementary S1. Transition probabilities assumption.**

**1. HR-HPV to LG-AIN**

Data on the Infected → LGAIN rate for HIV-positive MSM was not found in recent studies. Thus, we used data from a 1998 study by Palefsky et al. which measured AIN progression in 143 HIV positive and 131 HIV negative individuals for 2 years and also the number of HPV strains they were infected with [39]. The rate of progression in HIV positive individuals infected with either a single or multiple HIV strains was 0.58/person-year (95 cases in 165 person-years), and the rate of progression in HIV negative individuals was 0.30/person-year (32 cases in 106 person-years). Thus, the rate of Infected → LGAIN for HIV-positive MSM is 1.9 times that of HIV-negative MSM. The rates of progression in HIV negative individuals we estimated were similar to 0.078 [8], So the HIV positive MSM HR-HPV to LGAIN is 0.1482

**2. LR-HPV to GW**

LR-HPV infections were found in anogenital warts among MSM, with a prevalence of 75.9% in HIV-positive individuals and 41.7% in HIV-negative individuals [9]. This indicates that HIV-positive MSM have a higher risk of multiple LR-HPV infections compared to HIV-negative MSM, with a risk ratio of approximately 1.82. According to a MSM modelling study [10], the probability of LR-HPV to genital warts in MSM is 0.29. Therefore, for HIV-positive MSM, we assumed the probability of LR-HPV leading to GW is 0.29 * 1.82 = 0.5278.

**3. HG-AIN to Anal cancer**

**Annual transition probabilities from HG-AIN to anal cancer [11]**

| Age (years) | Mean |
|---|---|
| 12-35 | **0.0009** |
| 36-45 | **0.0014** |
| >45 | **0.0045** |

**4. Supplementary Table S5. Transition probabilities from HG-AIN to anal cancer among HIV- MSM.**

| MSM HIV- | | MSM HIV+ | |
|---|---|---|---|
| Age (years) | **Mean** | **Age (years)** | **Mean** |
| 12-35 | **0.0009** | **12-35** | **0.0099** |
| 36-45 | **0.0014** | **36-45** | **0.0154** |
| >45 | **0.0045** | **>45** | **0.0495** |

**Annual rates estimated from Machalek et al. [2], 1 in 377 in HIV-positive MSM and 1 in 4196 in HIV-negative MSM, with a risk ratio of 11:1. We assumed the annual rate for HIV-positive individuals is 11 times that of HIV-negative individuals.**

**5. LG-AIN to HG-AIN**

Data on the LGAIN → HGAIN rate for MSM by HIV status was not found in recent studies. Thus, we used data from a 1998 study by Palefsky et al. which measured incident HSIL (high-grade squamous intraepithelial lesion) in 277 HIV positive and 211 HIV negative individuals with 4 years of follow-up [39]. Of 135 HIV-positive MSM and 34 HIV-negative with LSIL (low-grade squamous intraepithelial lesion) or ASCUS (atypical squamous cells of undetermined significance) at baseline, 62 and 11 had HSIL by either cytology or histology within 4 years, for yearly rate of progression of 0.149 (416 person-years) and 0.096 (114 person-years), respectively. Thus, the rate of LGAIN → HGAIN for HIV-positive MSM is 1.5 times that of HIV-negative MSM. We use TreeAge Pro 2022 to transit the rate into probability by using function ratetoprob

**6. Supplementary Table S7. Anal cancer mortality among MSM stratified by age and HIV status.**

| MSM HIV- | | MSM HIV+ | |
|---|---|---|---|
| Age (years) | Mean | Age (years) | Mean |
| 12-29 | 0.0000 | 12-29 | 0.0000 |
| 30-39 | 0.0564 | 30-39 | 0.122952 |
| 40-49 | 0.2064 | 40-49 | 0.449952 |
| 50-59 | 0.1452 | 50-59 | 0.316536 |
| >59 | 0.2208 | >59 | 0.481344 |

The anal cancer mortality data for MSM who are HIV-negative comes from the Hong Kong Hospital Authority's local data. The death rate for HIV-positive individuals was found to be 2.18 times greater [5]. We assume that the death rate for every age group is consistently 2.18 times higher among MSM who are HIV-positive. Therefore, to estimate the mortality rates for HIV-positive individuals, we multiply the HIV-negative data by 2.18.

**7. Anal cancer to Cured**

The monthly transition probability for HIV-negative MSM from anal cancer to cancer cured is 0.0166, based on local data from the Hospital Authority. A retrospective cohort study found that the 5-year anal cancer-specific survival rates for HIV-negative and HIV-positive males are 80.5% and 72.5%, respectively [12]. The ratio of these survival rates is approximately 1:0.9. The cured rate of anal cancer among MSM is from [13]

**8. LR-HPV, HR-HPV to Susceptible**

Among people living with HIV, the HPV clearance rate is approximately halved, with a pooled relative risk (RR) of 0.53 (95% CI: 0.42 to 0.67) [14]. To adjust for the reduced clearance rate among HIV-positive individuals, we multiply the transition probabilities by the RR: For LR-HPV HR-HPV to Susceptible: 0.146 * 0.53 = 0.0774

**Supplementary Table S8. Number and Proportion of Anogenital Warts and Anal Cancer Cases Prevented under 9vHPV (70% Coverage)**

| | | | | Incidence of anogenital warts | Incidence of anal cancer |
|---|---|---|---|---|---|
| Base-case | | | | 1438.1 per 100,000 years | 2.03 per 100,000 person-years |
| Strategy | | | | Prevented anogenital warts | Prevented anal cancers |
| Age groups | | | | | |
| 12-18 | 19-27 | 28-44 | ≥45 | | |
| Strategies preventing the most anogenital warts | | | | | |
| One-group-targeted strategies | | | | | |
| 0 | 0 | 1 | 0 | 38330000 (26.7%) | 105500 (52.0%) |
| Two-group-targeted strategies | | | | | |
| 0 | 1 | 1 | 0 | 55750000 (38.8%) | 117700 (58.0%) |
| Three-group-targeted strategies | | | | | |
| 1 | 1 | 1 | 0 | 65800000 (45.8%) | 125900 (62%) |
| Strategies preventing the most anal cancer | | | | | |
| One-group-targeted strategies | | | | | |
| 0 | 0 | 1 | 0 | 38330000 (26.7%) | 105500 (52.0%) |
| Two-group-targeted strategies | | | | | |
| 0 | 1 | 1 | 0 | 55750000 (38.8%) | 117700 (58.0%) |
| Three-group-targeted strategies | | | | | |
| 0 | 1 | 1 | 1 | 65600000 (45.6%) | 138500 (68.2%) |
| Four-group-targeted strategies | | | | | |
| 1 | 1 | 1 | 1 | 75640000 (52.5%) | 143000 (70.4%) |

**Supplementary Table S9. The ICER values of different HPV vaccination strategies in sensitivity analyses.**

| | Strategy | | | | | | | |
|---|---|---|---|---|---|---|---|---|
| | All 12-18 | All 12-18 & 28-44 | All 12-44 | All ≥12 | HIV + 12-18 | HIV+ 12-18 & 28-44 | HIV+ 12-18 & ≥28 | HIV+ ≥12 |
| Vaccine price for both adolescent and adult | | | | | | | | |
| 25% | 580.4 | 6128.4 | 7522.6 | 8094.1 | Cost saving | Cost saving | Cost saving | 57.6 |
| -25% | Cost saving | 2255.9 | 3099.0 | 3966.0 | Cost saving | Cost saving | Cost saving | Cost saving |
| 50% | 1400.5 | 8064.6 | 9734.4 | 10418.8 | Cost saving | Cost saving | 322.5 | 1038.8 |
| Vaccine Efficacy Duration and time horizon | | | | | | | | |
| Protection of 10 years/ 20 year horizon | | | | undominated | | | | |
| Protection of 10 years/ 5 year horizon | 10465.6 | 24565.3 | 29466.9 | 31526.2 | Cost saving | 6631.0 | 7882.8 | 9717.4 |
| Protection of 20 years/ life time horizon | | | | undominated | | | | |
